# BCG VACCINES MAY NOT REDUCE COVID-19 MORTALITY RATES

**DOI:** 10.1101/2020.04.11.20062232

**Authors:** Samer Singh

## Abstract

The reason for the observed country-wise variability in incidence and severity of the COVID-19 outcome remains unknown. Few recent studies have suggested a positive protective correlation of the BCG vaccination policy of the countries with the observed COVID-19 severity. The current study was undertaken to reassess the existing data as of 4^th^ April 2020. The incidence rates (cases per million population), Case Fatality Rates (CFR) and inherently more robust Infection Fatality Rates (IFR) were calculated across countries accounting for about 99% COVID-19 deaths. The initial scrutiny suggested a weaker association with BCG vaccination policy or BCG coverage, so positivity to the Tuberculin Sensitivity Test (TST)/ Interferon Gamma Release Assay (IGRA) as a measure of the potential protective effect of the resident populations’ exposure to *Mycobacterium spp*. whether from BCG vaccination or as a result of exposure to environmental mycobacteria was analyzed. The incidence rates (the number of cases per million population) decreased with an increase in % LTBI (TST/IGRA positivity) for the analyzed countries with R^2 =^0.6343, suggesting an exponentially negative covariation. However, the covariation of CFR estimates that ranged from 0.29% to 12.25 % (average 5.39%) among countries, was tenuous. Interim estimates of IFR (i-IFR), a more dependable measure for such studies, for the best and worst-case scenarios, *i*.*e*., i-IFR-l and i-IFR-h, predict on an average 20.57% to 30.15 % COVID-19 fatality rates globally, but individual country estimates display huge variation. Among countries accounting for 92.14% deaths (11 countries; top 20% countries included in current study) the estimate for lowest IFRs (i-IFR-l=4.16 (China) & i-IFR-h=4.61 (China)) and highest IFRs (i-IFR-l=96.39% (UK); & i-IFR-h=96.54% (UK)) displayed huge difference (average for the group: CFR=6.8±3.6%; i-IFR-l=34.97±30.55%; & i-IFR-h=44.20±29.08%). Currently, the worst affected countries Italy (CFR=12.25%; i-IFR-l=42.63%; i-IFR-h=48.69%) and Spain (CFR=9.39%; i- IFR-l=26.85%; i-IFR-h=36.60%) would seemingly cope with COVID-19 better than UK, Netherlands and USA while the countries Germany (CFR=1.40%; i-IFR-l=4.93%; i-IFR-h=17.49%) and Switzerland (CFR=3.01%; i-IFR-l=10.87%; i-IFR-h=16.23%) along with China could fare the best. The rest of the 80% countries (accounting for 6.74% deaths), seemed to have reduced mortality (CFR=2.45±2.01; i-IFR-l= 30.62±28.24%; i-IFR-h=40.99±30.47%) with associated high % LTBI (17.28±8.87) than top 20% countries. The inherent issues in the data set (e.g., heterogeneity, non-random sampling, different criteria of sampling and reporting, access to health care, genetic composition, underlying co-morbidities, etc) need to be taken into account for making informed decisions.

## Introduction

Current COVID-19 pandemic caused by SARS-CoV-2, a coronavirus closely related to SARS and MERS, had a humble beginning in late 2019 in Wuhan, China. It rapidly spread to the majority of the nations by mid-March 2019. The COVID-19 pandemic has stirred worldwide havoc with higher infection rates and associated variable adverse outcomes across countries. So far the most affected countries had been the developed West and South European, North American countries along with China and Iran from Asian countries. It had already killed about 60 thousand people by 4^th^ April 2020 when the analysis being communicated was planned [1]. With exponentially increasing deaths every passing day, the pandemic has sent the countries and their health services scrambling to decrease the fallout of COVID-19. The dedicated worldwide platforms had been created to provide up-to-date information about the pandemic. The overall recovery rate for COVID-19 had been about 79% making all countries panic to consolidate all resources to decrease the deaths. As of today, 11^th^ April 2020 the death ascribed to COVID-19 has already crossed the 100 thousand mark [1]. Various clinical trials to repurpose the existing drugs indicated for similar coronavirus or suggested pathology, as well as experimental vaccines, have been initiated/planned [2,3]. A large number of studies are also being conducted and results published in journals or posted online on preprint servers medRxiv and bioRxiv to understand the basis of differential incidence and fatality rates in different countries. Few studies indicate the existence of a positive correlation between BCG vaccination policies to the inherent resistance of populations to COVID-19 and suggest clinical trials to evaluate its preventive potential in COVID-19 [4,5]. The data available needs to be analyzed thoroughly to arrive at actionable context-specific conclusions to ameliorate the devastating effect of COVID-19 on human lives.

The bacille Calmette–Guérin (BCG) vaccine is primarily given to protect against tuberculosis (TB) in countries with higher TB incidence [6]. TB remains the top cause of death from a single pathogen *Mycobacterium tuberculosis* (*MTB*). The vaccination of BCG or exposure to environmental non-tuberculous mycobacteria is supposed to ‘train’ cell-mediated immunity to respond better on subsequent exposure to intracellular pathogen MTB by responding through CD4- and CD8- positive cells and the production of various cytokines including Interferon-γ (IFN-γ) which is also a key cytokine in innate and adaptive immunity against viral and bacterial infections [6,7]. A more generalized long-term cross-reactive protective effect against several pathogens is suggested through long-term epigenetic programming and ‘training’ of immune cells particularly Macrophages and NK-cells by this mycobacterial exposure (BCG or environmental). The BCG vaccination has been found to provide protective immunity for a variable duration in different populations ranging from 15 years to 50-60 years [8,9]. However, it should be noted that in about 90% of the cases MTB gets cleared even in the absence of BCG vaccination or prior ‘training’ of the immune system. BCG vaccination does not prevent primary infection of MTB, activation of latent TB infection (LTBI) and supposed to have only a limited effect on the prevention of MTB spread in a population [10]. The supposed protective immunity wanes away with time but subsequent exposures to environmental mycobacteria could act as a booster to keep the immunity intact. The exposure to *Mycobacterium spp*., whether it is MTB, BCG vaccine or environmental isolate, is usually tested by Tuberculin Sensitivity Test (TST) and Interferon Gamma Release Assay (IGRA).

The current analysis was conducted to ascertain the effect of the prevalence of immune response against *Mycobacterium spp*. in select populations to their susceptibility to COVID-19. It intends to provide evidence to make an informed decision by the policymakers.

## Material and Methods

The data about coronavirus pandemic was collected from the website https://www.worldometers.info/coronavirus/. The analysis utilizes data about BCG vaccination, coverage in different years, TB incidence and latency collected by Institute for Health Metrics and Evaluation (IHME), 2018 Global Burden of Disease Collaborative Network. Global Burden of Disease Study 2017 (GBD 2017) Results. Seattle, United States and WHO data presentation collected from https://ourworldindata.org/coronavirus-data [11]. The Microsoft excel was used for basic descriptive data analysis.

## Results and Discussion

Recently a correlation, though weak, had been suggested between universal BCG vaccination implementation and its potential protective role in the occurrence of lower COVID-19 cases and consequent death as a result of COVID-19 in some countries. We had accessed the COVID-19 pandemic data on April 4^th^, 2020 (Updated on April 04, 2020, 00:55 GMT) for different countries from website https://www.worldometers.info/coronavirus/ (See supplementary file: sheet ‘Table 1 COVID- 19 CASES 04 April’. Data of countries reporting more than 1000 cases that included countries with universal BCG vaccination policy and good current BCG coverage but supposedly less severe COVID 19 impact, like India, Brazil, etc. were selected for further analysis. These countries accounted for about 99% of total deaths and about 98% of total COVID 19 cases (See supplementary file: sheet ‘Top 20- bottom 80% contributors’).

A visual comparison of the country-wise confirmed COVID-19 death rate per million people as on 4^th^ April 2020 to universal BCG vaccination policy, actual estimated BCG vaccination coverage, Healthcare Access and Quality (HAQI) Index, Neglected tropical diseases (NTDs) and Tuberculosis incidence are presented in Figure 1. When we consider countries contributing to the majority of the cases with an incidence rate of 25 to >200 cases per million, most of them appear in the higher bracket of economies with HQI index of more than 70 (see Fig 1A with 1B.) Apparently, better healthcare and hygiene are puzzlingly translating into a higher incidence of COVID-19. Recently, it has been proposed that the earlier implementation of universal BCG policy could be responsible for the lower incidence of COVID-19 [4]. BCG vaccine is generally given to children in high TB prevalence countries to protect them from developing pulmonary TB. Its vaccination is estimated to protect different populations for variable durations ranging from 15 to 60 years [8,9]. The BCG induced delayed T-cell mediated hypersensitivity is supposed to protect through training the immune system to respond in a more disciplined manner without getting overwhelmed with ‘cytokine storm’ when exposed to pathogens [7,12]. When we compared the COVID-19 incidence among different countries with BCG vaccination policy implementation (Figure 1C) and the total BCG coverage (Figure 1 D), they do not seem to affect the COVID-19 confirmed deaths per million population for specific countries (e.g., Australia, USA, Germany). Countries falling in less than 89% BCG coverage regions seemingly display more disparate outcomes concerning coronavirus deaths (e.g., Finland, Sweden, South Africa, Iran, Greenland, Iceland, etc).

**Figure 1.**
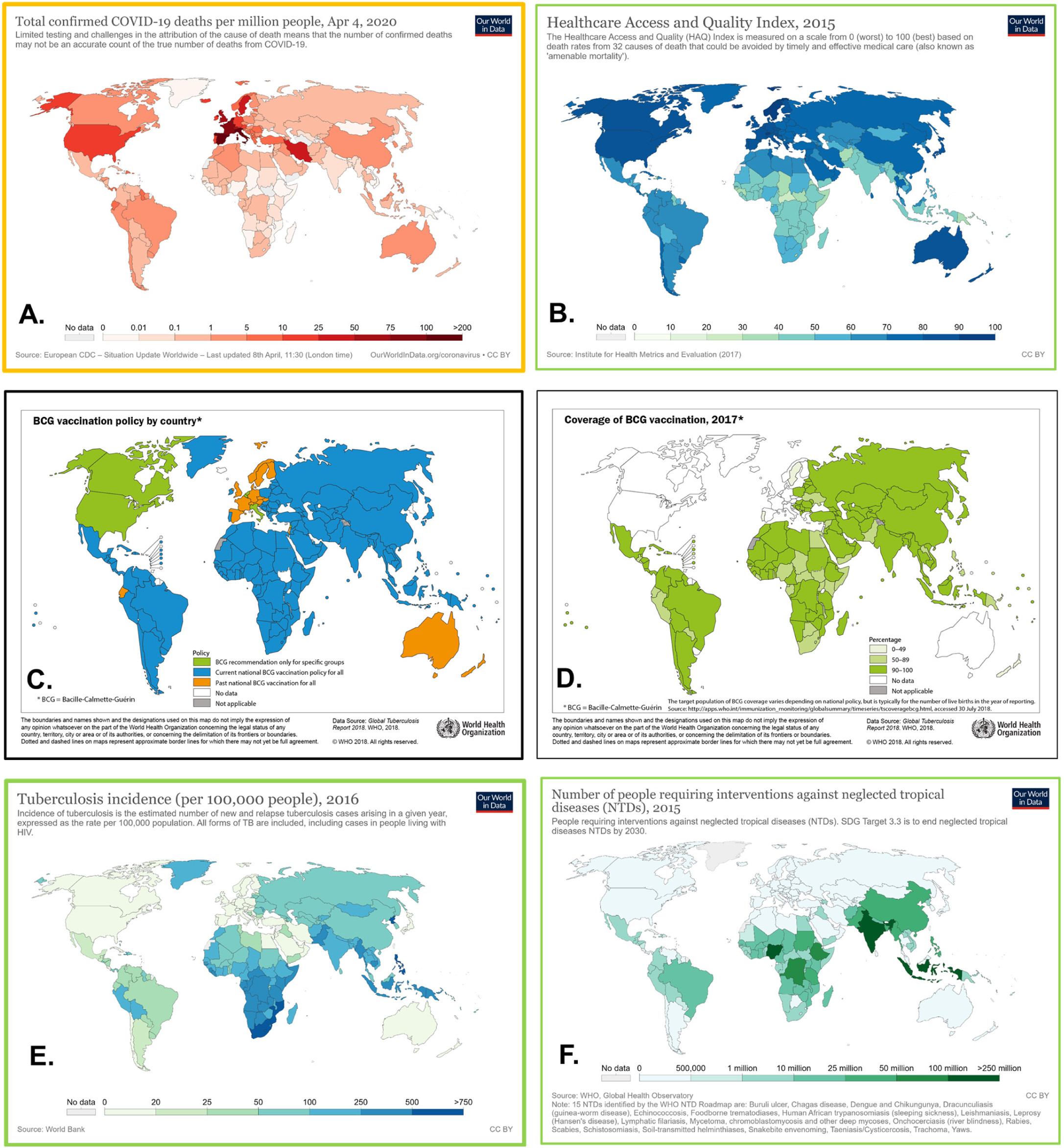
BCG vaccination policy implementation and total coverage does not seem to affect the COVID-19 confirmed deaths per million population in different countries. The observed total COVID-19 confirmed deaths per million of population among different countries (A) seem to correlate positively with Health Access and Quality index (B) while negatively to Tuberculosis incidence (E) and Neglected Tropical disease (NTDs) burden of the population (F). Its dependence on BCG Vaccination policy implementation (C) and more importantly coverage of BCG vaccination (D) appears much weaker. Disclaimer: Images A, B, E and F are from https://ourworldindata.org/coronavirus while C and D are from https://www.who.int/imaqes/default-source/maps/qlobal_tb_beg_vaccination_policy_2O17.pnq?sfvrsn=9ff3babf_0 for illustrative purpose. The copyright of the image rests with the entities disclosed. They are being used for non-commercial purpose as fair use entity.

We hypothesized, maybe the overall burden/incidence of TB and Neglected Tropical diseases (NTDs) may display a better correlation with deaths from COVID-19. When we compared the COVID-19 incidence with TB or NTD burden of the population, the COVD-19 seems to affect more the countries with lower disease burden especially with TB Overall, the observed total COVID-19 confirmed deaths per million of population among different countries seem to correlate (negatively) more with Tuberculosis incidence than coverage of BCG vaccination or BCG vaccination policy implementation as expected if these disease burden may provide some nonspecific protection against COVID-19.

We reasoned if there is some nonspecific protective immunity at work against COVID-19 due to *Mycobacterium spp*. exposure of the resident population (BCG vaccination or environmental isolates) the occurrence of COVID-19 cases in a population would correlate better with LTBI percentage estimated for different countries. We wondered if the rate of LTBI (%) (reported based on TST/IGRA positivity), among different countries (total 55 countries with more than 1000 cases) without any regard for income, healthcare facilities, BCG vaccination policies or BCG coverage, would show a relationship with the numbers of COVID-19 cases per million population. Surprisingly, the number of cases per million showed a decline with an increase in % LTBI (TST/IGRA positivity) with R^2 =^0.6343, suggesting an exponentially negative covariation between the two or in other words higher TST/IGRA positivity could be protecting against COVID-19 or contributing to a slower spread of the COVID-19 as compared to countries with lower TST/IGRA positivity (Figure 2 A). As inherent with any statistical analysis for correlation, the observed correlation cannot be construed as to indicate a cause and effect relationship by default. A more simple explanation for the observed correlation could be the higher frequency of people’s movement between the countries currently having a high burden of COVID-19 and the country of COVID-19 origin. The most affected countries of the West and China, for being prominent tourist destinations and economic workhorses, had been inherently exchanging a large flux of visitors.

**Figure 2.**
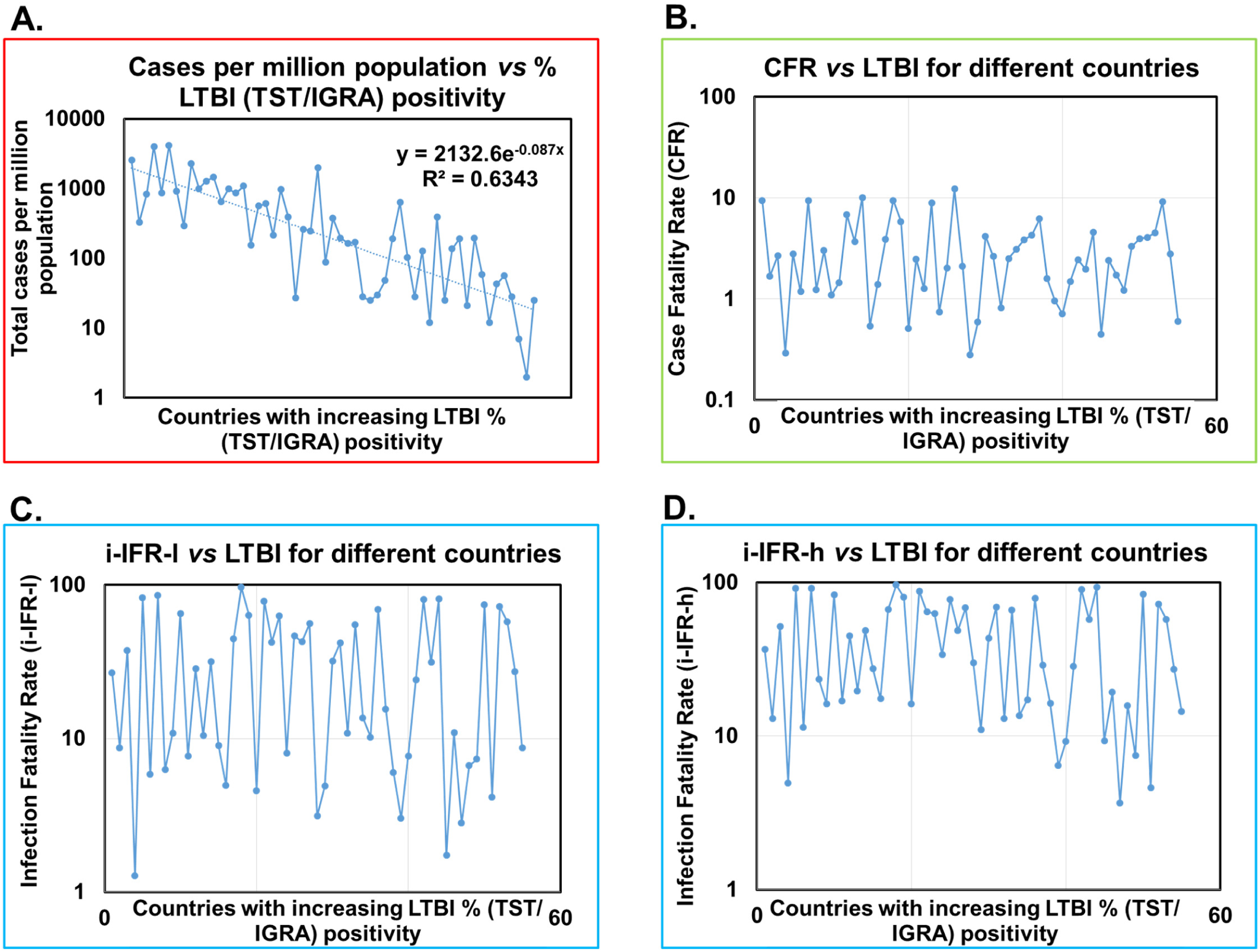
The Case Fatality Ratio for COVID-19 does not correlate with %TST/IGRA positivity of populations. **(A)** The incidence of COVID-19 cases per million population decrease with increase in %LTBI or TST/IGRA positivity (left to right) among countries reporting >1000 cases. **(B)** Case Fatality Rate varied from 0.29 to 12.25% among countries without depending on TST/IGRA positivity. **(C) & (D)** Estimated interim Infection Fatality Rate (i-IFR) lower (i-IFR-l; 1.28 to 96.39%) and higher (i-IFR-h; 3.66 to 96.54%) also seem to not correlate with TST/IGRA positivity among countries. Note: Most countries on right side are in an earlier stage of pandemic.

Case Fatality Rate/ Ratio (CFR) is an estimate of how probable it is that someone (case) may die after contracting a disease. CFR estimates are known to vary by location, phase of the outbreak, screening criteria adopted, the extent of screening, reporting issues –number of cases identified/reported, supporting health care, etc. As the COVID-19 pandemic is still underway, the true CFR cannot be reliably ascertained, rather just guessed to allow us to have some understanding of its evolving epidemiology. The CFR value for COVID-19 had been recently estimated between 0.2% (Germany) to 7.7% (Italy) [13]. To evaluate whether BCG vaccination may be providing a certain degree of protection (correlation again!) against COVID-19 to people in countries reporting high BCG coverage (high TST/IGRA positivity), we calculated the CFR in the same set of countries reporting more than 1000 cases, using definition *CFR* (*in %*) *=* [*Total deaths from disease / Total number of cases reported*] *x 100* and plotted them vs % LTBI (TST/IGRA) positivity (Figure 2B). The CFR values ranged from 0.29% to 12.25 % among countries. The countries with lower %LTBI (<13% (6.07- 12.97%); total 27) seem to have higher CFR than next 28 countries with higher %LTBI (>13% (13.25- 47.64%). However, it should be noted that these lower CFR displaying countries (higher %LTBI) on an average are also supposed to be relatively at an earlier phase of the pandemic (except China) and accounting for <15 % of total deaths (including China) at the moment. Though a negative correlation can be obtained, it would be tenuous.

To get a more realistic actionable estimate of the current emerging COVID-19 situation for the projection purpose, we had estimated *interim* - *Infection Fatality Rate/ Ratio* (*i-IFR*) using definition *i- IFR* (*in %*) *=* [*Total deaths from disease/ Total number of cases with outcomes*] × 100. These estimates are required to assess the gravity of the situation. The lowest possible IFR (i-IFR-l) calculation, for the best possible outcome scenario, included both dead and recovered for the total number of cases with outcomes. The highest possible IFR (i-IFR-h) calculation, for the worst possible outcome scenario, considered the critically sick in the dead category (as a possibility) and rest was similar to i-IFR-l calculation. This was done to get a more realistic estimate of the problem for two reasons: currently, we do not have a complete picture of COVID-19’s course and an estimate from the Chinese outbreak projects a period of 2 to 8 weeks from the appearance of first symptoms to death [14]. The predicted/projected death rate for COVID-19 under the best possible scenario (i-IFR-l), when all critically sick will recover, ranged from 1.28% (for Iceland) to 96.39% (For UK) while for the worst-case scenario estimate (i-IFR-h), when all critically sick may die, varied from 3.66 % (for S. Korea) to 96.54% (for UK) in (See Supplementary File sheet: Table for Figure 2). It is interesting to note that the difference observed in the CFR, i-IFR-l and i-IFR-h estimates are quite surprising across countries. The CFR to i-IFR-l increase was lowest, less than 1%, for China (4.07-4.61%) while it witnessed more than 80 % increase for UK (9.44 to 96.54%) and Netherlands (9.45% to 91.83%). Every countries context is different and those variables need to be identified to better manage the COVID-19. However, when i-IFR-l and i-IFR-h estimates were plotted vs increasing % LTBI positivity of different countries’ populations, no apparent correlative inference could be made (Figure 2C and 2D) about the % LTBI positivity of countries and the severity (IFR) of the COVID-19.

The large variation observed in the estimates of COVID-19 infection fatality rate, i.e., i-IFR-l and i- IFR-h, for different countries (about 5% to almost 97 %) cannot be credibly explained with available data. We speculate this large variation could be the result of countries being in different stages of the pandemic as well as inherent differences in the affected population demography, co-morbidities and genetic makeup as also indicated for select populations such as South Korea, Spain, China, Italy [11,14]. Detailed comparative analysis of the personal health records of patients with final outcomes (*i*.*e*., dead, recovered, sick) along with genetics if available need to be performed wherever possible in all affected countries to better understand the underlying confounding variables affecting COVID- 19 outcomes and also design a better strategy to manage it.

Next, for getting a better picture of the underlying mechanics if any concerning role of % LTBI of the target population, we divided the selected countries into top 20 % (11 countries; each accounting for >500 death) that accounted for 92.14% of total COVID-19 deaths and bottom 80% (44 countries) accounting for approximately 6.74 % of the COVID-19 deaths. Analysis of case fatality rate (CFR) and interim Infection fatality rate (i-IFR) for countries was made (Supplementary File: Sheet ‘Top 20- bottom 80% contributors’ and Figure 3). The calculated CFR and i-IFR values for the top 20% countries are presented in tabular form as Figure 3A (for the bottom 80% contributors refer Supplementary File: Sheet ‘Top 20- bottom 80% contributors’). Though higher TST/IGRA positivity (LTBI) could be argued to cause lower total COVID 19 cases per million (compare high %LTBI countries Iran and China with other nations with lower %LTBI population), the death rate per million population comparison seems to make the association murkier. A closer examination of the table indicates against any possible broadly applicable protective contribution of high % TST/IGRA positivity (LTBI) to overall CFRs for COVID-19 in respective countries. For example countries with lower %LTBI and no BCG vaccination policy or coverage such as Germany (CFR=1.40%), USA (CFR=2.67%) and Switzerland (CFR=3.01%) have their CFRs lower than China (CFR=4.07%) or Iran (CFR=6.19%) which have both BCG policy and coverage in place as well as higher %LTBI. Surprisingly countries which are seemingly worst-hit now with regard to total number no of deaths, will seemingly have lower IFRs (i-IFR and i-IFR-h), e.g., Italy (CFR: 12.25%; i-IFR-l=42.63%; i-IFR- h=48.69%), Spain (CFR: 9.39%; i-IFR-l=26.86%; i-IFR-h=36.60%) as compared to many others who currently seem to be less severely affected, e.g., UK (CFR: 9.45%; i-IFR-l=96.39%; i-IFR-h=96.54%), Netherlands (CFR= 9.46%; i-IFR-l=85.61%; i-IFR-h= 91.83%). Current combined estimates for these countries stand as CFR=6.8±3.9, i-IFR-l=34.97±30.55%; i-IFR-h=44.20±29.08%. Next, we compared these countries’ fatality rate estimates with that of the bottom 80% countries (44 countries; accounted for 6.74 % COVID-19 deaths) and presented the comparison as a bar diagram (Figure 3B). The error bars indicate STDEV. Though average CFR and i-IFR estimates are lower for the bottom 80% countries (CFR=2.45±2.01; i-IFR-l=30.62±28.24%; i-IFR-h=40.99±30.47%) with associated high % LTBI (17.28±8.87) as compared to that for top 20% countries, any definitive correlation may not be appropriate concerning %LTBI (TST/IGRA positivity) due to large variations among the population of different countries (Supplementary File: Sheet ‘Top 20- bottom 80% contributors’) unless country-specific discounts are made to exclude some. It did not escape our attention that different groupings of countries can still be made to show a possible positive correlation between LTBI (TST/IGRA positivity) percentages of the country population with lower COVID-19 severity.

**Figure 3A.**
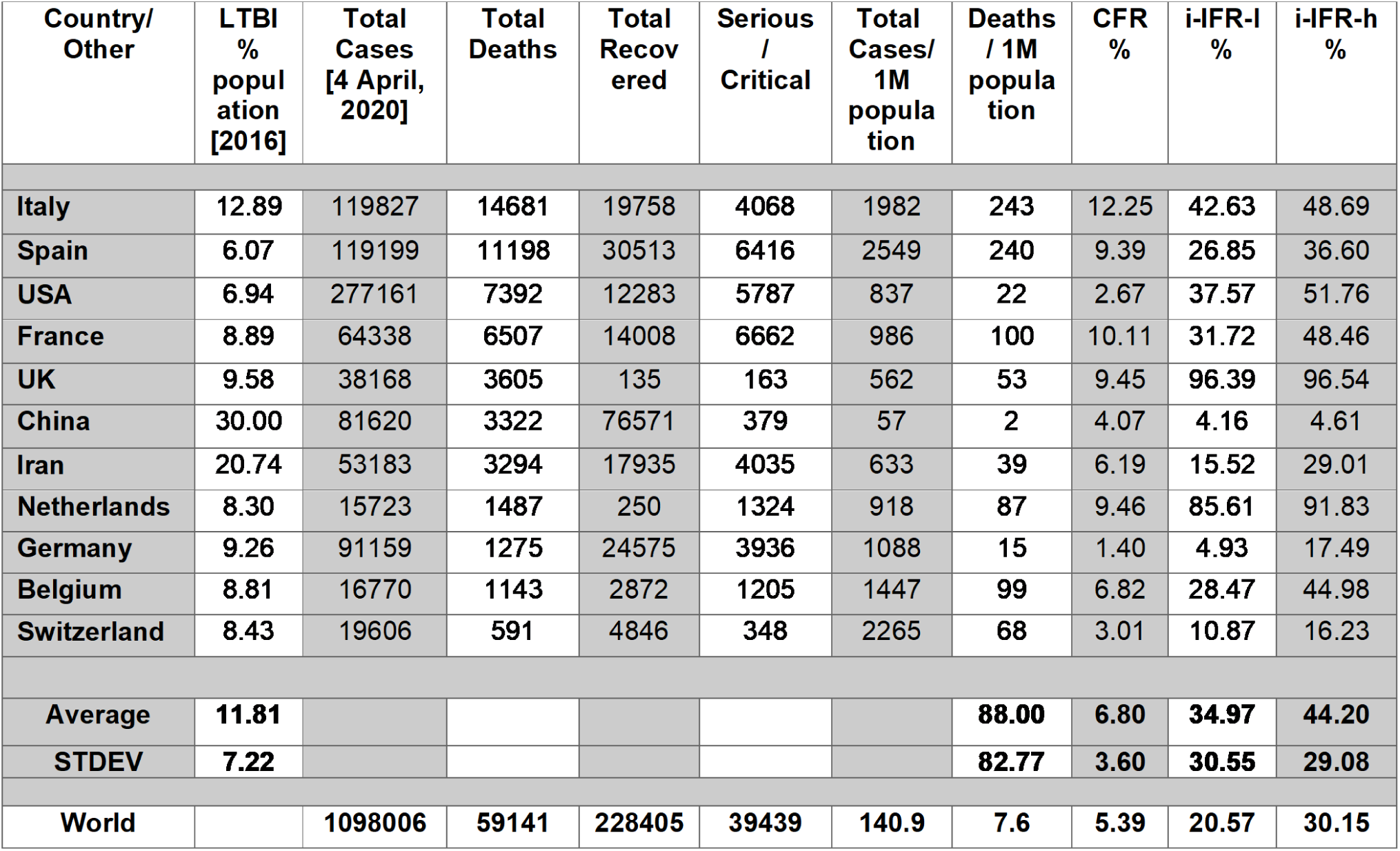
Analysis of CFR and IFR for countries contributing 92.14 % of total COVID-19 deaths (top 20 % countries included in the study with >500 cases)

**Figure 3B.**
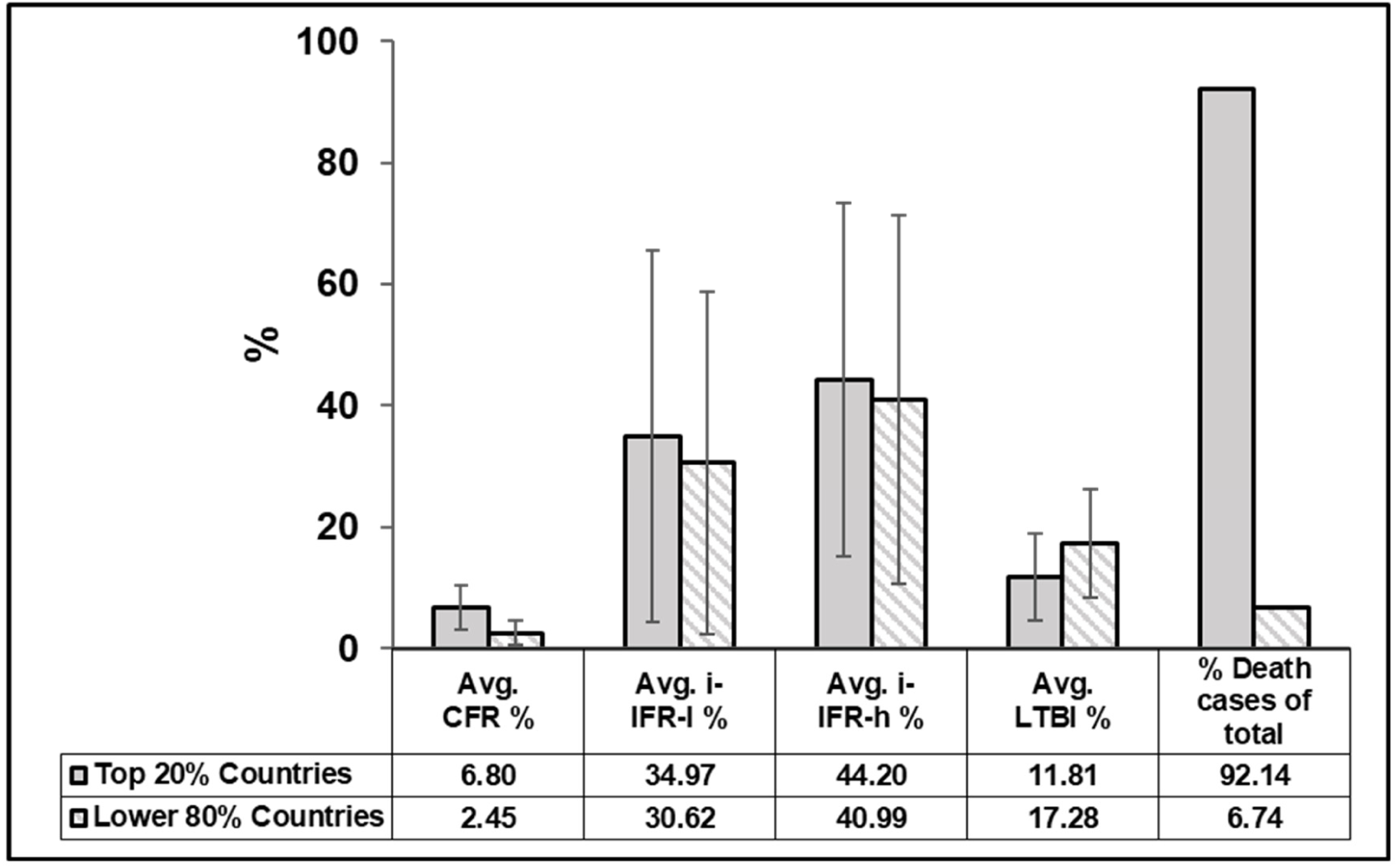
The CFR and IFR variation between top 20% and bottom 80% countries (contributing total 98.9 % deaths) included in the study.

There is an urgent need to identify the risk factors which may be promoting the COVID-19 adverse outcomes. Some risk variables such as age, cardiovascular disease, chronic respiratory disease, diabetes have been already identified as associated with adverse outcome in select populations, e.g., Italy, South Korea, Spain, China *etc* [11,14]. These potential causative association needs to be verified in other populations as well. The underlying mechanisms responsible for adverse outcome needs to be elucidated and the causes precisely identified for taking any corrective measures to decrease the overall COVID-19 mortality. It would be also interesting to investigate the underlying issues with the reporting, data collection or identify protective contributory factors which may be responsible for such disparate outcomes. Difference in the virulence of virus strain currently affecting different population and the ‘training status’ of immune system to deal with intracellular pathogens (exposure to mycobacteria/BCG/TST positivity) to mount measured immune response without buckling under the ‘cytokine storm’ as predicted for SARS-CoV-2 infection, could be some of the other risk factors for select populations/groups.

## Conclusion

The BCG vaccination may not be the primary cause for the lower infection and mortality rate per million population observed in select countries who had adopted the universal BCG policy early on or covering the entire population for vaccination. Though the idea that the exposure to environmental Mycobacterium spp. or BCG vaccine could train the body to respond to intracellular pathogens better cannot be completely discounted but the marginal benefit, if any, which could be supposedly gained would be limited to specific population subgroup susceptible to developing TB.

There are countries with hugely disparate case fatality rates among both low % LTBI or BCG coverage countries (High: Italy, Spain, USA etc. *vs* Low: Germany, Switzerland, Austria, Iceland) and high %LTBI or BCG coverage countries (High: Brazil, Philippines, Indonesia, etc *vs* Low: Japan, China, Mexico, S. Africa, S. Korea, Thailand etc.). The large variation in CFR and more importantly in estimated interim IFRs among countries could be resulting from the inherent population constitution, e.g., genetics, age, underlying co-morbidities (e.g., cardiovascular disease, diabetes, age, HIV status, inflammatory disease like rheumatoid arthritis) or due to different inherent virulence/pathogenicity of SARS-CoV-2 virus strains currently circulating in different regions. The underlying causes of the difference in the fatality rates among different country populations need to be understood. The government agencies may collect and make available more patient specific data pertaining to COVID- 19 to allow its closer scrutiny for making better informed decisions. For correlative analysis, Immune Marker Variation/ HLA typing and genetic background information may be already available or readily obtained in the developed countries who are also currently some of the most affected countries with the COVID-19. The studies exploring the correlation of the genetic makeup of the COVID-19 patients (*i*.*e*., critical, recovered, deceased, along with asymptomatic and those displaying non-serious symptoms) may be actively promoted or taken up on a priority basis. It is time for various international bodies, government agencies and philanthropists to come forward and allow this kind of retrospective/prospective work to be undertaken to better understand the adversary SARS-CoV-2019. Current situation, warrants the research organizations and groups with required resources to be brought together to work in unison towards the common goal of containing the COVID-19 spread and devastation on a war footing as the window of opportunity may be closing at a rate faster than perceived.

## Data Availability

All data referred is publically available and also included in the submission

## Acknowledgement

The funding support from Banaras Hindu University to the laboratory of SS is acknowledged. The author gratefully acknowledges the fruitful constructive discussions on the subject with Professor Rakesh Bhatnagar, Jawaharlal Nehru University, New Delhi, India that helped it take the current form.

## Disclaimer

*Images in Figure 1: A, B, E and F are from https://ourworldindata.org/coronavirus while C and D are from https://www.who.int/images/default-source/maps/global_tb_bcg_vaccination_policy_2017.png?sfvrsn=9ff3babf_0 for illustrative purpose. The copyright of the image rests with the respective entities. They are being used here for non-commercial purpose as fair use entity. Wherever applicable, permission from the copyholders may be sought for their commercial reproduction*).

## Notes

### Competing Interest Statement

The authors have declared no competing interest.

### Funding Statement

No funding was received for the submitted work.

